# A generalized SEIRD model with implicit social distancing mechanism: a Bayesian approach for the identification of the spread of COVID-19 with applications in Brazil and Rio de Janeiro state

**DOI:** 10.1101/2020.05.30.20117283

**Authors:** Diego T. Volpatto, Anna Claudia M. Resende, Lucas dos Anjos, João V. O. Silva, Claudia M. Dias, Regina C. Almeida, Sandra M. C. Malta

## Abstract

We develop a generalized SEIRD model considering social distancing measures to describe the spread of COVID-19 with applications in Brazil. We assume uncertain scenarios with limited testing capacity, lack of reliable data, under-reporting of cases, and restricted testing policy. We developed a Bayesian framework for the identification of model parameters and uncertainty quantification of the model outcomes. A sensitivity analysis is performed to identify the most significant parameters on either the cumulative numbers of confirmed and death, or the effective reproduction number. We show the model parameter related to social distancing measures is one of the most influential. Different relaxation strategies of social distancing measures are then investigated to determine which strategies are viable and less hazardous to the population. The considered scenario of abrupt social distancing relaxation implemented after the occurrence of the peak of positively diagnosed cases can prolong the epidemic, with a significant increase of the projected numbers of confirmed and death cases. A worse scenario occur if the social distancing relaxation policy is implemented before evidence of the epidemiological control, indicating the importance of the proper choice of when to start relaxing social distancing measures. The employed approach and subsequent analysis applied over the Brazilian scenarios may be used to other locations.

## 1. Introduction

At the end of 2019, the world was taken by the news about the outbreak in China of a new coronavirus called SARS-CoV-2, which stands for *Severe Acute Respiratory Syndrome Coronavirus 2*. The associated disease, called COVID-19 by the World Health Organization (WHO), rapidly spread around the world and is currently considered a pandemic. As of September 30, 2020 there have been 235 countries and territories affected, with around 33.8 million confirmed cases and 1 million confirmed deaths [1]. Coronavirus belongs to a group of viruses that are common in humans and are responsible for 15% to 30% of cases of common cold [2]. The SARS-CoV-2 has spread rapidly and has already affected a significant part of the world’s population [3]. The within-host dynamics seem to be very heterogeneous and the severity of the disease varies widely. Based on currently available information, elderly people and people of any age with comorbidities, such as hypertension, cardiovascular disease, and diabetes, are among those at most risk of severe illness from COVID-19. In most locations, the burden of COVID-19 cases requiring medical intensive care exceeds the available medical resources, worsening the situation. A great effort has been made worldwide to better understand the disease and possible mechanisms to control it. Measures such as widespread testing, and reducing social contacts for some age segments, or for the wider population, are known to decrease the speed of the disease spread and the case fatality [4]. However, countries often have to deal with limitation of testing capacity for COVID-19 which increases the uncertainty associated with how the disease behaves and is likely to complicate the management of the policies to mitigate and control it. One form to deal with this lack of information is through mathematical modeling, using diversified modeling techniques (see [5] for a review). This type of modeling also has some limitations, because it intrinsically relies on a simplification of reality. One example of such simplification is the usual assumption of constant parameters values (see [6] for an overview of applications and limitations of mathematical modeling for COVID-19). Population-based models that split individuals into classes are widely used, and is the approach followed in this work.

WHO has already recognized that testing for COVID-19 is a key way to know how the virus spread and to provide insights on how to respond to it, although widespread testing is low in most countries in the world. In Brazil (BR), for example, the initial testing policy included only severely ill and healthcare practitioners people. The sub-notification of infected cases was, and still is, a major problem, coupled with incomplete and inconsistent overall data as well as the lack of complete knowledge on the prevalence and progression of the disease [7]. Likewise, the number of deaths may be underestimated for the same reason. Overall, one of the main difficulties in developing predictive compartmental models of the COVID-19 is the lack of reliable data to support parameter choices. In the face of the current scenario, we developed a seven-compartment model that implicitly considers the social distancing policy that isolates individuals from the infection for a period of time. This modeling strategy to consider social distancing is also employed in [8] using a generalization of the standard SIR model. Here we focus on understanding the model response to perturbations/uncertainties. We perform sensitivity analysis and quantify the uncertainties in model outcomes, mainly the cumulative numbers of confirmed and death cases, and the effective reproduction number. We also investigate hypothetical scenarios on how to obtain a desired effect/control via a modification to the system subject to uncertainties. Our studied cases include the whole Brazilian scenario (BR) and the state of Rio de Janeiro (RJ), which was one of the first states in BR to recommend and implement social distancing measures, beginning on March 17, 2020 [9]. However, they have not last much longer and RJ has started a progressive social distancing flexibilization, as the whole country [10].

## 2. Mathematical Modeling and Methods

### 2.1. The SEAIRPD-Q Model

We develop a generalized Susceptible–Exposed–Infected–Removed–Dead (SEIRD) model that includes protective social distancing measures based on the following assumptions: (i) the analysis time is small enough such that natural birth and death are disregarded; (ii) all positively diagnosed individuals, who are severely ill, are hospitalized; (iii) only symptomatic infected and positively tested (diagnosed/hospitalized) individuals may die due to complications from the disease; (iv) the hospitalized individuals are under treatment and remain isolated, so that they are considered not infectious; (v) social distancing measures are restrictive so that isolated individuals are not likely to be infected, and (vi) the recovered/removed individuals acquire immunity. Our SEAIRPD-Q model has seven compartments, including the population of positively diagnosed (*P*) individuals who are under medical treatment, and the infected class that is modeled as two separate compartments encompassing individuals with and without symptoms, denoted by *I* and *A*, respectively. The removed compartment (*R*) includes the recovered individuals as well as those under social distancing measures. The mathematical description (differential equations) of the SEAIRPD-Q model and its schematic representation are shown in Fig. 1 while model parameters and related meanings are exhibited in Tables 1 and SM-A.1 (see the Supplementary Material (SM) for more details).

**Table 1:**
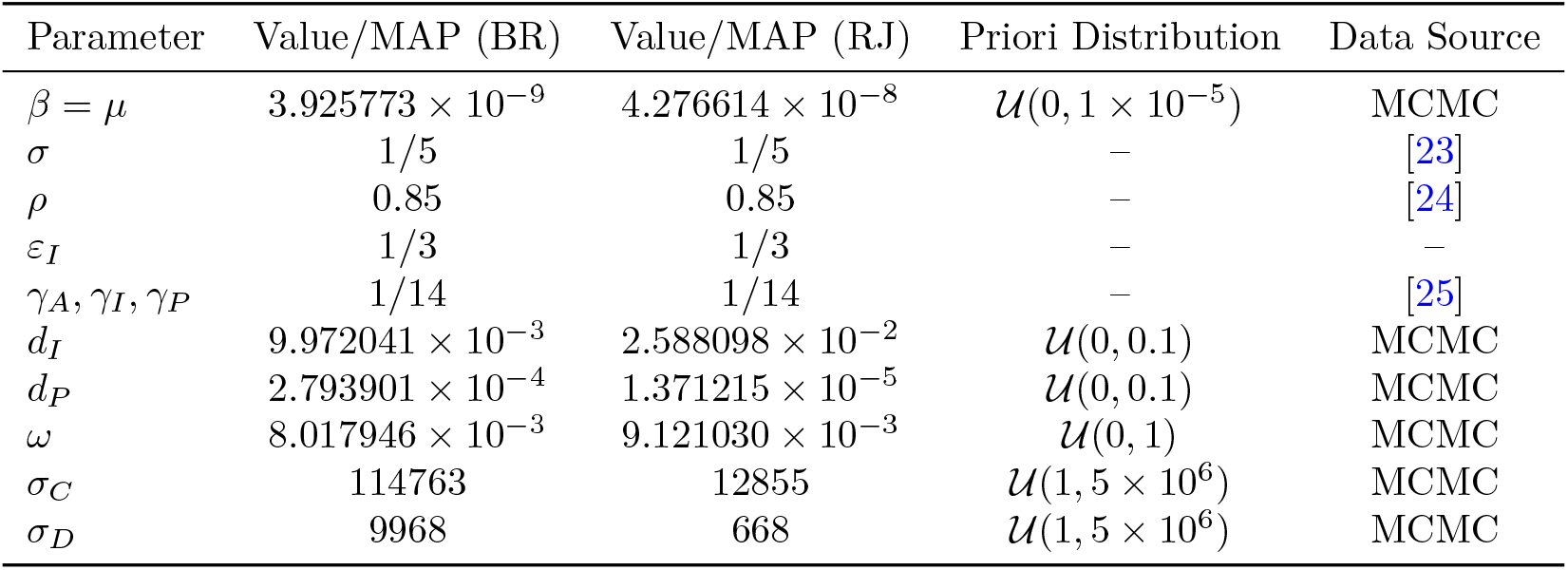
Model parameters: fixed and MAP estimates for BR and RJ (in appropriate units). A more detailed description of all variables, parameters, and ICs are shown in the SM.

| Parameter | Value/MAP (BR) | Value/MAP (RJ) | Priori Distribution | Data Source |
| --- | --- | --- | --- | --- |
| $\beta = \mu$ | $3.925773 \times 10^{-9}$ | $4.276614 \times 10^{-8}$ | $\mathcal{U}(0, 1 \times 10^{-5})$ | MCMC |
| $\sigma$ | 1/5 | 1/5 | — | [23] |
| $\rho$ | 0.85 | 0.85 | — | [24] |
| $\varepsilon_I$ | 1/3 | 1/3 | — | — |
| $\gamma_A, \gamma_I, \gamma_P$ | 1/14 | 1/14 | — | [25] |
| $d_I$ | $9.972041 \times 10^{-3}$ | $2.588098 \times 10^{-2}$ | $\mathcal{U}(0, 0.1)$ | MCMC |
| $d_P$ | $2.793901 \times 10^{-4}$ | $1.371215 \times 10^{-5}$ | $\mathcal{U}(0, 0.1)$ | MCMC |
| $\omega$ | $8.017946 \times 10^{-3}$ | $9.121030 \times 10^{-3}$ | $\mathcal{U}(0, 1)$ | MCMC |
| $\sigma_C$ | 114763 | 12855 | $\mathcal{U}(1, 5 \times 10^6)$ | MCMC |
| $\sigma_D$ | 9968 | 668 | $\mathcal{U}(1, 5 \times 10^6)$ | MCMC |

**Figure 1:**
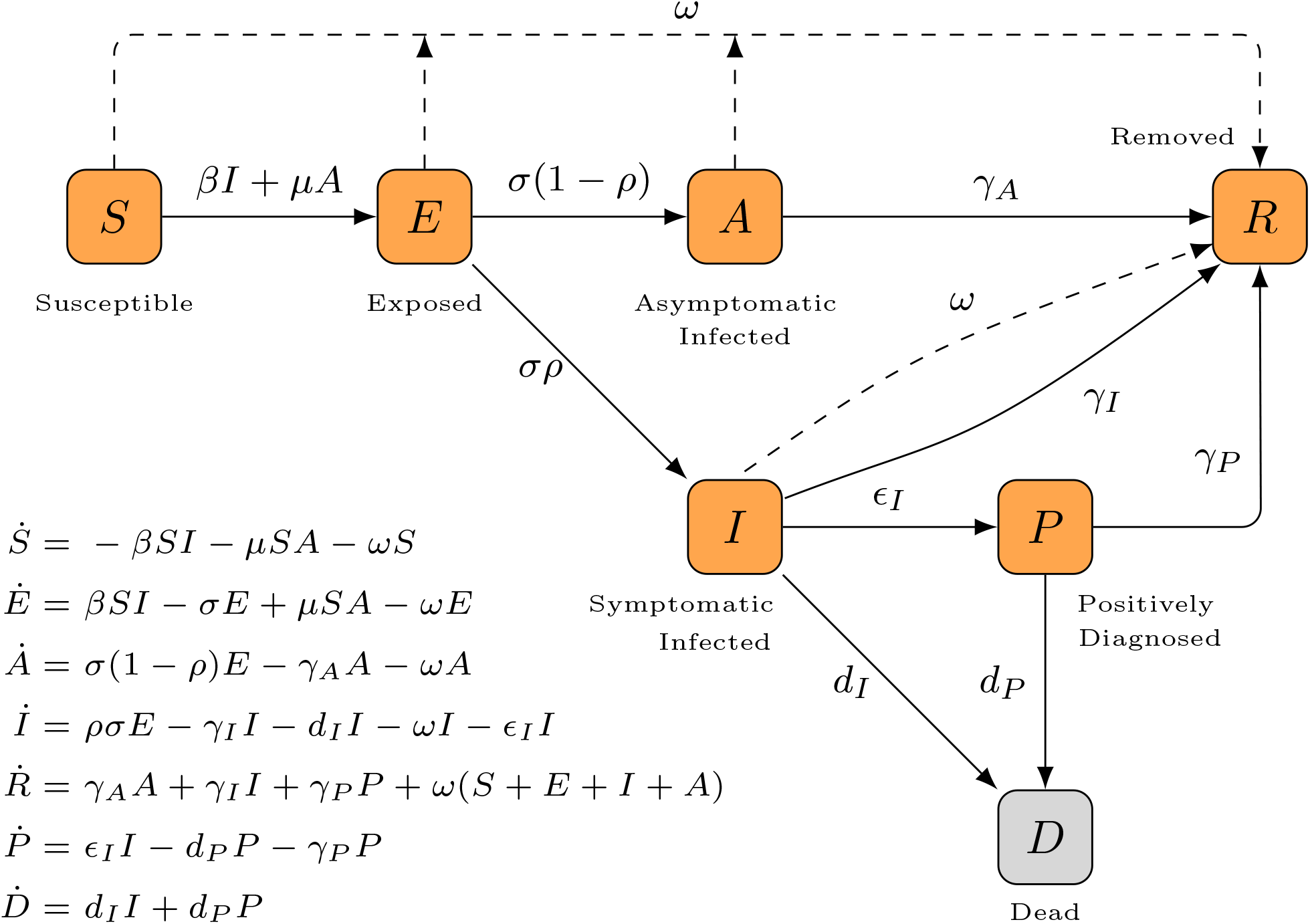
Mathematical formulation and schematic description of SEAIRPD-Q model.

Susceptible individuals become exposed to COVID-19 when in contact with infected people. The rate at which this happens may vary depending on whether the infected individual has symptoms or not, denoted by *β* or *μ*, respectively. After an incubation period (1*/σ*), exposed individuals become infected. The fraction *ρ* of infected people who present symptoms can vary widely. Due to the assumption of reduced testing capacity, asymptomatic individuals, as well as symptomatic infected individuals with mild symptoms, are not likely to be hospitalized and therefore will not be diagnosed. Those who present symptoms are either isolated or hospitalized. We assume that only symptomatic infected individuals with stronger symptoms are diagnosed quickly, at a rate of *ϵ*_*I*_, and require hospitalization. This assumption better reflects the policy on only testing severely ill individuals. Following the WHO guideline on social distancing, most countries around the world are adopting social distancing policies at some extent. An interesting feature of our model is the assumption that susceptible, exposed, and infected individuals can be kept isolated at a removal rate of *ω*, an assumption also taken by [8]. Thus, the removed individuals’ compartment includes individuals who have recovered from the disease as well as those subjected to social isolation. Asymptomatic and symptomatic individuals can recover without medical treatment at rates of *γ*_*A*_ and *γ*_*I*_, respectively, and hospitalized individuals recover at a rate of *γ*_*P*_. The cumulative numbers of confirmed and death cases are obtained, respectively, from simulation day zero (*t*_0_) to a desired time *t* as:

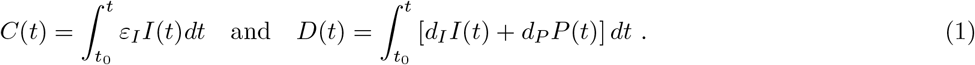

### 2.2. Evaluation of the Reproduction Number

Following the ideas introduced in [11], we apply the Next Generation Matrix method to obtain ℛ_0_ for the SEAIRPD-Q model. Firstly, we use a reduction process to arrive at a linearized infection model, under the disease-free steady state. The identification of the infected variables (*E, A*, and *I*) allows defining the vector **x**^T^ = [*E, I, A*], where the superscript T stands for the transposition of a matrix. Thus, we consider that *S, R, P*, and *D* individuals are not able to transmit the disease. Denoting by *T* and Σ the transmission and transition matrices, respectively, the three-dimensional linearized infection sub-system is given by:

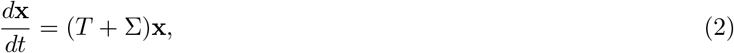

where

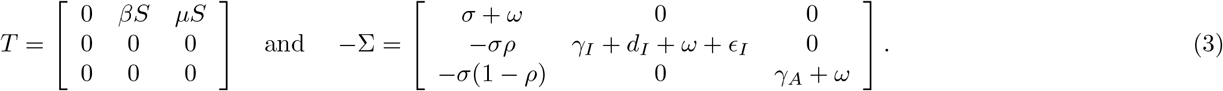

The reproduction number *ℛ*_0_ is the dominant eigenvalue of the next generation matrix *K* = *−T* Σ^−1^ [11], which yields:

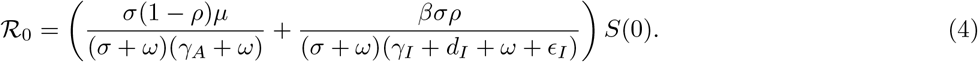

The corresponding effective reproduction number is:

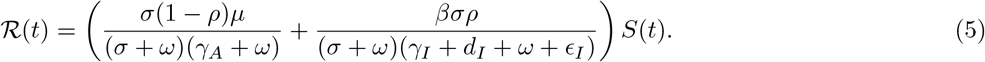

### 2.3. Bayesian Calibration

Model calibration is performed using a Bayesian approach to make results consistent with available observations on cumulative confirmed and death cases. With Bayesian calibration, we are capable of determining the most likely uncertainties for input parameters as well as their maximum *a posteriori* (MAP) estimates of marginal distributions described by the *a posteriori* probability distribution *π*_post_. Considering that our aim is to adjust a set of parameters ***θ*** given ***y*** observations, Bayes’ theorem states:

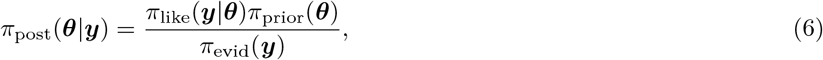

where *π*_prior_ is the *a priori* probability distribution that represents the initial (*a priori*) knowledge over ***θ***, *π*_like_ is the likelihood function, and *π*_evid_ is the evidence (information) encompassed in the data (see [12] for a more detailed description). We assume a Gaussian likelihood function in the form:

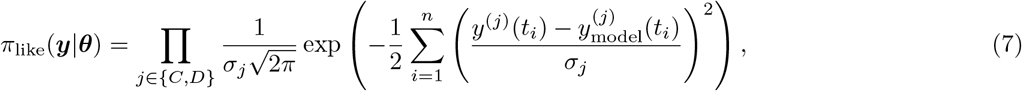

where *y*^(*C*)^(*t*) and *y*^(*D*)^(*t*) are the observable quantities of cumulative numbers of confirmed and death cases, respectively, and their corresponding simulated outcomes are 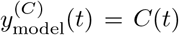 and 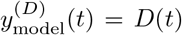. Gaussian noise is associated with each observable quantity with variances 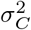 and 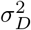, which are considered hyperparameters to be determined in the Bayesian calibration procedure.

Due to paucity data and possible model identifiability issues, we calibrate the following model parameters: *β* = *μ, ω, d*_*I*_, and *d*_*P*_. All other parameters are gathered from the available literature and listed in Table SM-A.1 (see SM for more details).

The code implementation is written in Python language (version 3.7), using PyMC3 as a Probabilistic Programming framework [13]. Due to the intensive computational burden, we choose as calibration method the Cascading Adaptive Transitional Metropolis in Parallel [14], a Transitional Markov chain Monte Carlo available in PyMC3. For the sake of reproducibility, code and scripts are publicly provided at [15]. Of note, the SEAIRPD-Q system of seven ordinary differential equations is solved using the LSODA method [16] from SciPy [17]. This package provides an interface for such routine from ODEPACK [18].

### 2.4. Data

We investigate the model behavior for BR and RJ. We gathered data for *C* and *D* from [19]. Since Brazilian policy for testing in COVID-19 is still mostly restricted to severe cases, model calibration aims at matching *C* and *D*, without accounting the recovered outflow from *P*. We assessed the epidemiological data of 198 days for BR, ranging from March 5, 2020, to September 18, 2020, and 193 days for RJ, ranging from March 10, 2020, to September 18, 2020. The initial dates were chosen considering a minimal requirement of at least five diagnosed individuals at the pandemic initial date. For completeness, all used data are listed in [15].

### 2.5. Sensitivity Analysis

In order to quantify how changes in the model parameters and some initial conditions (ICs) affect the quantities of interest (QoI), we apply the Elementary Effects method [20] present in the SALib library [21]. In contrast to local sensitivity methods, which evaluate how small changes around a singular point affect a QoI [22], the Elementary Effects method is a global sensitivity method. Here we analyze QoI_1_(*t*) = *ℛ* (*t*) and the normalized sum of the squares of *C* and 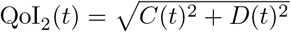. Considering ***θ*** *∈* ℝ*k* as the set of analyzed parameters, we construct a sample of *m* initial points ***θ***^(1)^, …, ***θ***^(*m*)^ from a *k*-dimensional *p*-level grid. Afterwards, a metric defined as an elementary effect *EE*_*i*_, *i* = 1, …, *k*, is computed for every parameter:

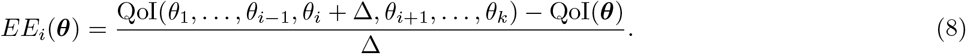

where 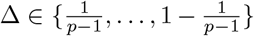 is the magnitude of change in the *i*th parameter in the *k*-dimensional *p*-level grid. Values of *p* as an even number and 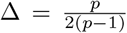 are usual suggested choices to ensure an equal sampling probability in the parametric space, and are the default option in the SALib library [21]. After computing the elementary effects, the process ends by calculating the global sensitivity indexes for each parameter. Our work dedicates the analysis only to the first order sensitivity index:

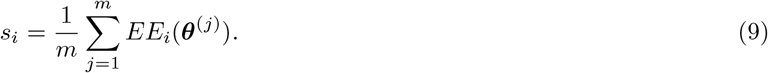

Parameters with high scores are the most influential to the QoI. In turn, the first order sensitivity index values allow ranking the parameters with respect to their order of importance.

### 2.6. Time Dependent Social Distancing Policy Modeling

To represent and understand social distancing policy effects on COVID-19 epidemic, we propose a time-dependent removal rate, which we denote by *ω*_*r*_. Considering that *t*_*d*_ is the time at which the relaxation policy is implemented, *ω*_*r*_ is defined as the following continuous function:

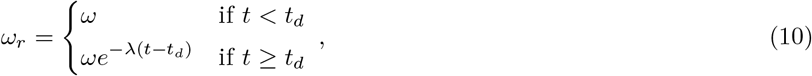

in which *λ* = ln 2(*t*_1/2_)^−1^ is the decay constant, with *t*_1/2_ being the half-life time for the social distancing release policy. In this way, *ω*_*r*_ is a smooth decreasing function after *t* = *t*_*d*_ for which *λ* regulates the decay speed.

## 3. Results

To properly perform projections and further study different social distancing scenarios of interest, we calibrated the SEAIRPD-Q model to the data of BR and RJ (separately). We employed a Bayesian calibration of some model parameter values, with remaining parameter values gathered from the literature. Model parameters are shown in Table 1, while the frequency histograms for the calibrated parameter posterior marginals and other model factors are presented in the SM.

Taking into account the maintenance of the social distancing policies during the time of the analysis, Fig. 2 exhibits the fitting and predictions for the COVID-19 pandemic in BR. Fig. 2a shows that the peak of *P* individuals occurred on the simulation day 145 (95% CI: 143–146) with around 638.8 thousand (95% CI: 630.6–647.1) people simultaneously infected (denoted “active cases”, hereinafter). By the end of the simulation time, the model predicts around 149.3 thousand (95% CI: 145.1–153.6) of deaths and 4.392 million (95% CI: 4.306–4.482) of cumulative cases. The ℛ_0_ calculated by means of MAP for BR is 3.09, and the time evolution of ℛ(*t*) is displayed in Fig. 2b (black line). Notice that ℛ (*t*) *<* 1 occurred around the simulation day 135 (95% CI: 133–136), which indicates that the disease is controlled [26].

**Figure 2:**
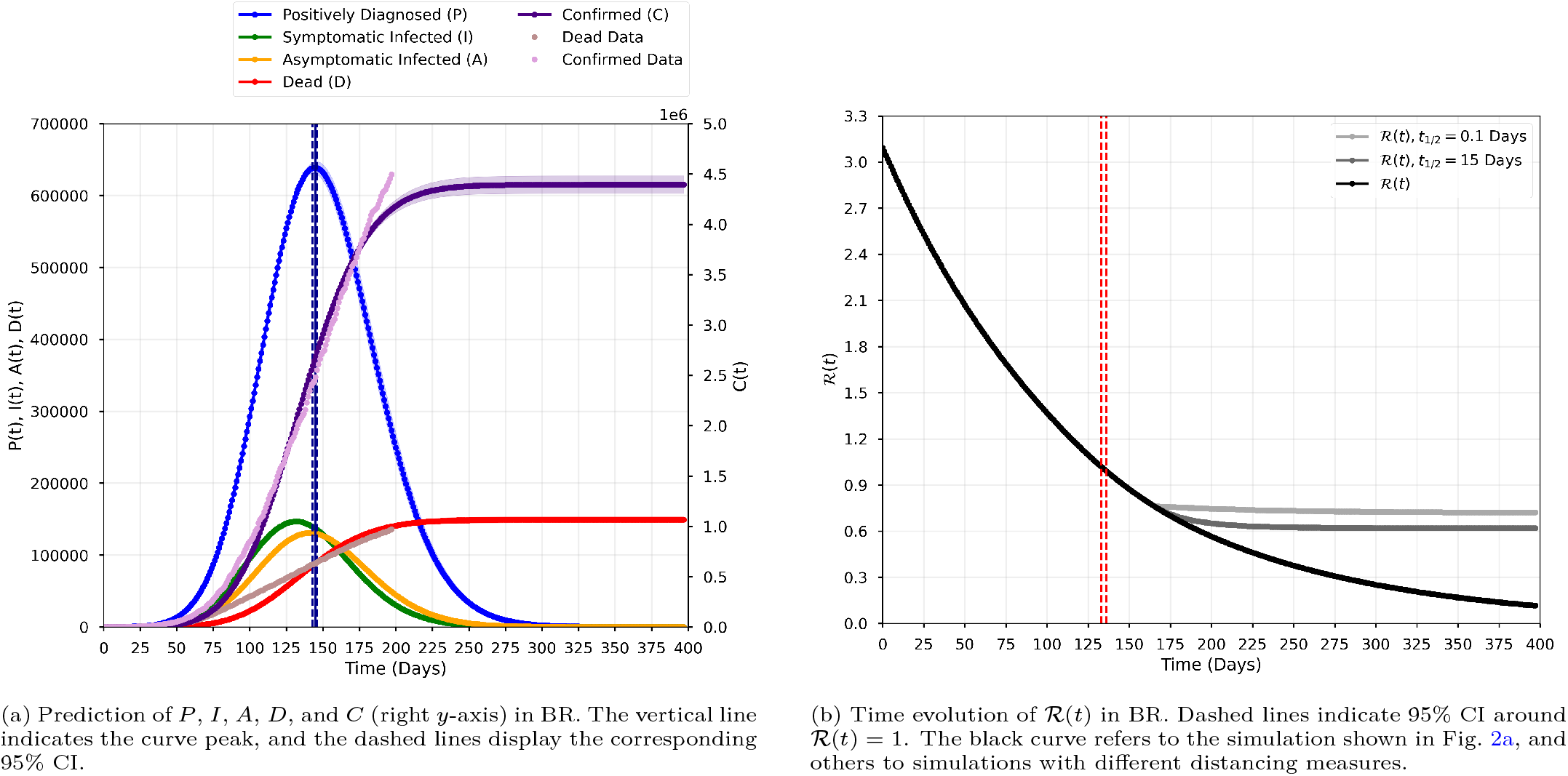
Dynamics of the COVID-19 in BR modeled with the available data. The lines indicate the simulation with the MAP estimates, with the shaded colors indicating the 95% credible interval (CI). Simulation days 0 and 145 (peak day of active cases) corresponds to March 5, 2020 and July 28, 2020, respectively.

We also fitted the model with the available data for RJ. Considering again the maintenance of the social distancing policy, the projections for COVID-19 infection in RJ are shown in Fig. 3. Of note, model fitting for RJ is much worse than in the BR scenario, probably driven by highly noisy data. Nevertheless, the model could capture correctly the peak of active cases, that occurred on the simulation day 115 (95% CI: 114–116) with around 32.2 thousand (95% CI: 31.7–32.7) active cases. Also, around 16.6 thousand (95% CI: 16.3–17.0) of deaths and 212.5 thousand (95% CI: 207.6– 217.7) of cumulative cases are expected on day 225, when these numbers stabilized. The ℛ_0_ is 2.68 for RJ, and the time evolution of ℛ (*t*) is displayed in Fig. 3b (black line). Similarly as for the broader scale of the Brazilian scenario, if the social distancing measures are maintained in RJ, the disease was under control around the simulation day 105 (95% CI: 103–107), when the effective reproduction number becomes less than one.

**Figure 3:**
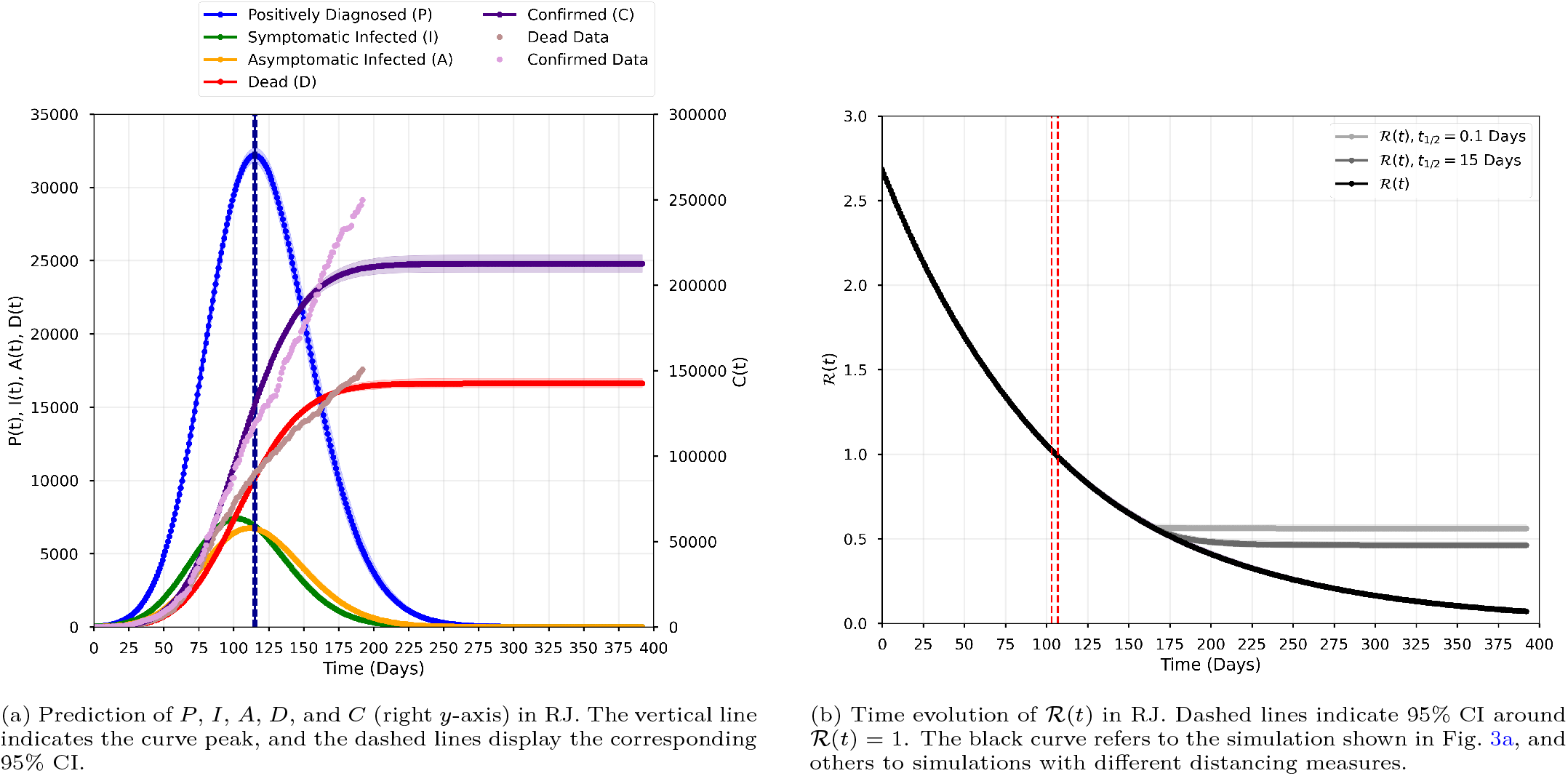
Dynamics of the COVID-19 in RJ modeled with the available data. The lines indicate the simulation with the MAP estimates, with the shaded colors indicating the 95% CI. Simulation days 0 and 115 (peak day of active cases) corresponds to March 10, 2020 and July 3, 2020, respectively.

We address the sensitivity for both *C* and *D* (QoI_2_(*t*)), as well for ℛ (*t*) (QoI_1_(*t*)). Sensitivity analysis is performed for both scenarios (BR and RJ) considering all parameters and the initial conditions *E*(0), *A*(0), and *I*(0). The settings *p* = 4 and *m* = 40 are used for all experiments. A 50% variation around the fixed and MAP values in Table 1 is considered to construct parameter ranges.

The sensitivity analysis results for BR are depicted in Fig. 4. The sensitivity indexes for the cumulative *C* and *D* (QoI_2_(*t*)) are heavily influenced by the initial conditions in the early stages of the disease propagation, although their influence decreases as time evolves. In contrast, they are not influential for ℛ (*t*) (QoI_1_(*t*)) along all the simulation time. The influence of the parameter related to social distancing (*ω*) on both QoIs is remarkable. More importantly, its influence increases with the evolution of time, which emphasizes the need for care when establishing social distancing relaxation measures. On the other hand, the proportion of infected individuals who have symptoms is also one of the most influential parameters on both QoIs, although it is more relevant considering *C* and *D*. This points out the need of having a more widespread testing policy. Regarding the RJ scenario, Fig. 5 reveals very similar scores. One noteworthy distinction is the rank of influence of *E*(0) and *I*(0) associated with QoI_2_(*t*). The former is more influential in the BR scenario (Fig. 4b), while the latter has a higher score for RJ (Fig. 5b). Overall, the most influential parameter is the removal rate (*ω*), and its impact increases as the dynamics evolves. The second most influential parameter is the proportion of *I* with symptoms (*ρ*). We remark that the latter parameter is fixed due to the limited testing capacity in BR.

**Figure 4:**
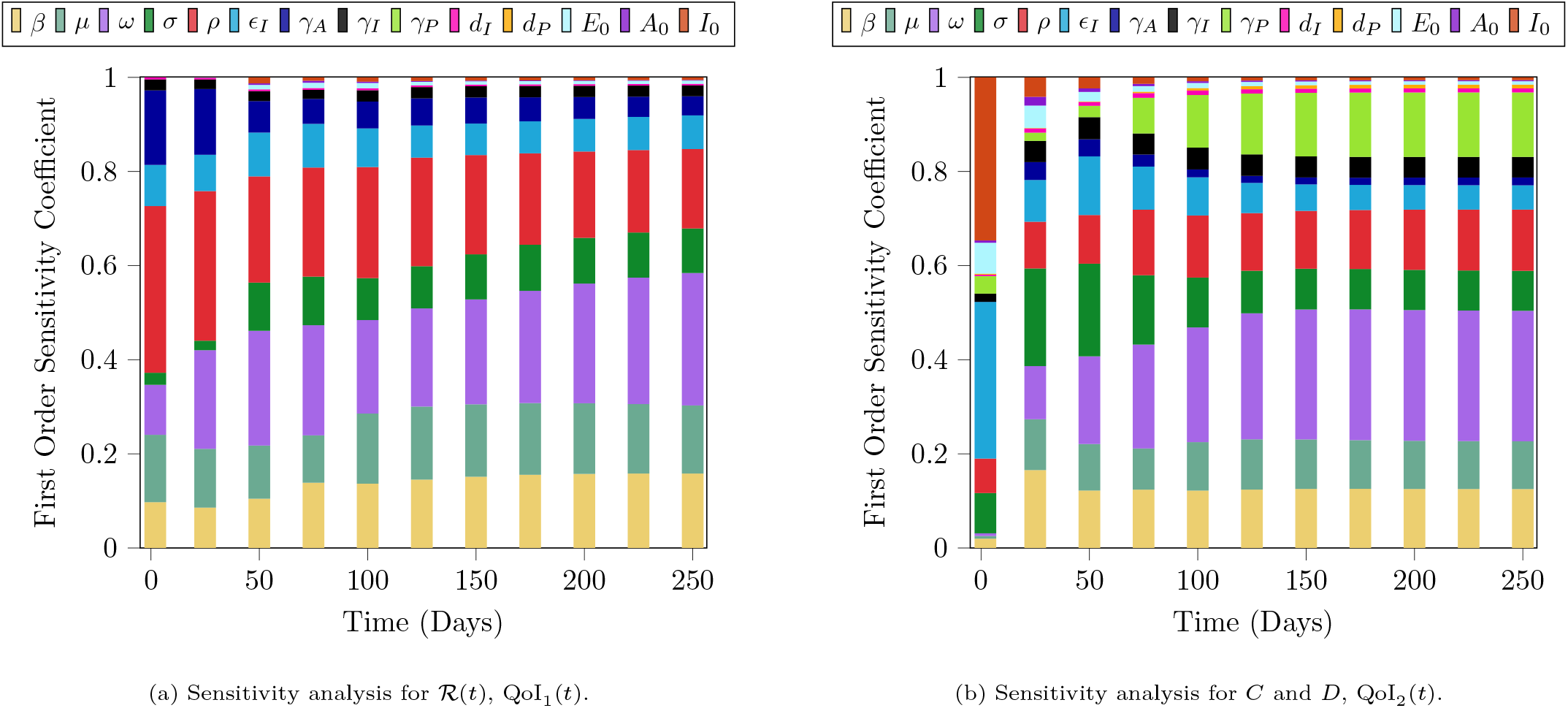
Temporal changes of the first order sensitivity index of model factors for the BR scenario.

**Figure 5:**
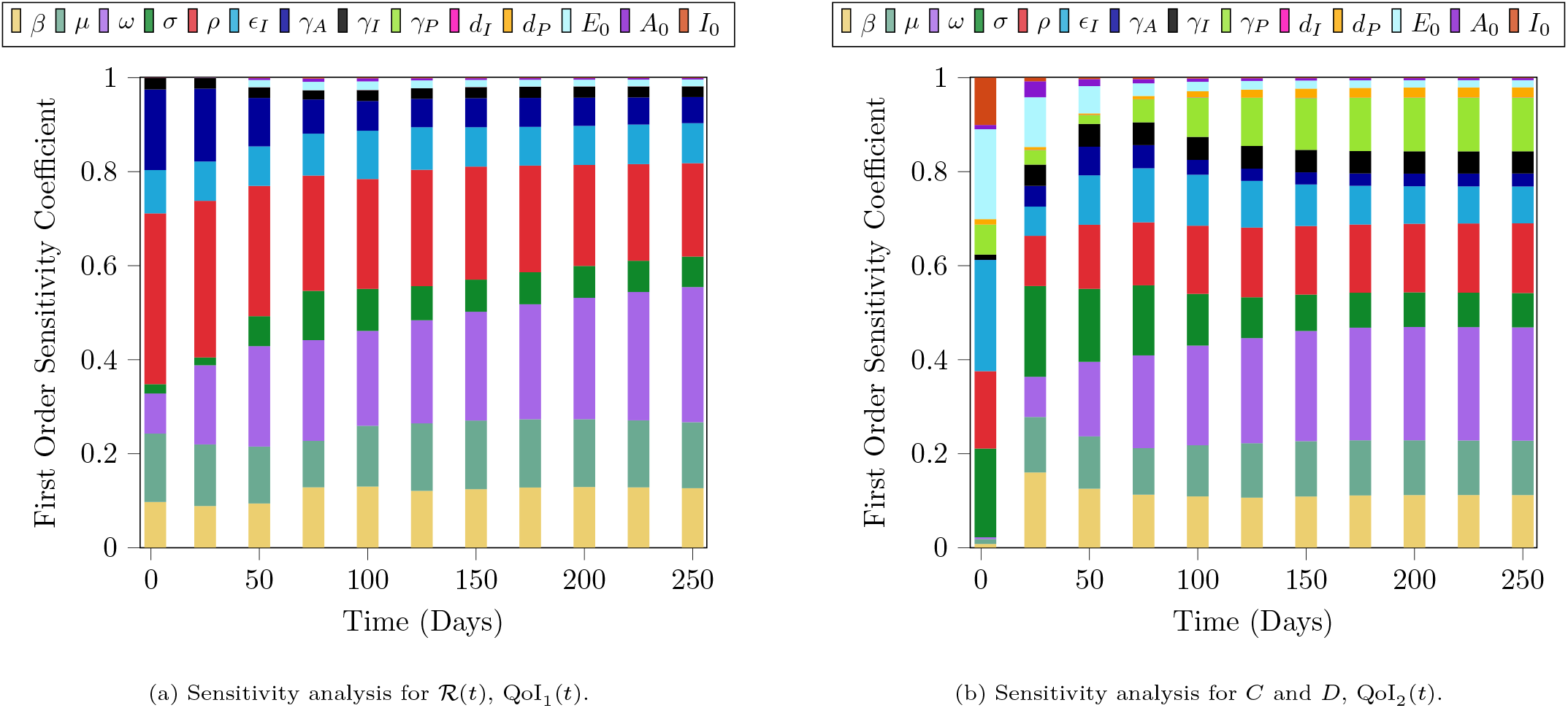
Temporal changes of the first order sensitivity index of model factors for the RJ scenario.

### 3.1. A study case: How would different social distancing measures affect the disease spread?

We have shown that the rate at which *S, E, A*, and *I* are removed due to social distancing measures significantly affect ℛ(*t*), *C*(*t*), and *D*(*t*). Such average values under the actual social distancing policy are an idealization of reality since actual values are dynamic and spatially heterogeneous. However, such idealization is useful to qualitatively analyze the effect of different disease control mechanisms. Thus, in order to study the qualitative effects of changes in social distancing policies, we propose to model *ω* as defined in (10), which can represent different social distancing relaxation scenarios. We consider three cases: (i) a sudden release from social distancing, which is modeled as a decay with half-life as *t*_1/2_ = 0.1 days; (ii) a gradual release, with half-life as *t*_1/2_ = 15 days, and (iii) the current social distancing policy, with constant removal rate. The latter represents the original study case, which can also be obtained by setting *t*_*d*_ *>* 400 days. For cases (i) and (ii), we selected *t*_*d*_ = 165 days, time for which the peak of active cases has already passed. Fig. 6 shows the consequences of relaxing social distance measures in terms of the model distributions of *C* and *D* for BR at *t* = 397 days. Additional results are displayed in the SM for both BR and RJ scenarios. Remarkably, a sudden release can induce an increase in *C* and *D*, and their uncertainties, extending the crisis duration due to a slower decrease in the active cases (see the SM for more details). This effect can also be noted in Figs. 2b and 3b, where we show the time evolution of ℛ(*t*) for cases (i) and (ii) in comparison with the original scenario. Social distancing release policies make the decrease of ℛ(*t*) far slower, implying in a slow control of the disease. Thus, determining an appropriate moment to begin a social distancing release policy demands special care, since applying such policies in inappropriate times can maintain a crisis status for a longer-term. Moreover, due to the small changes in ℛ (*t*) for a longer time, more cases of *C* and *D* will occur, worsening the health damage in the total population.

**Figure 6:**
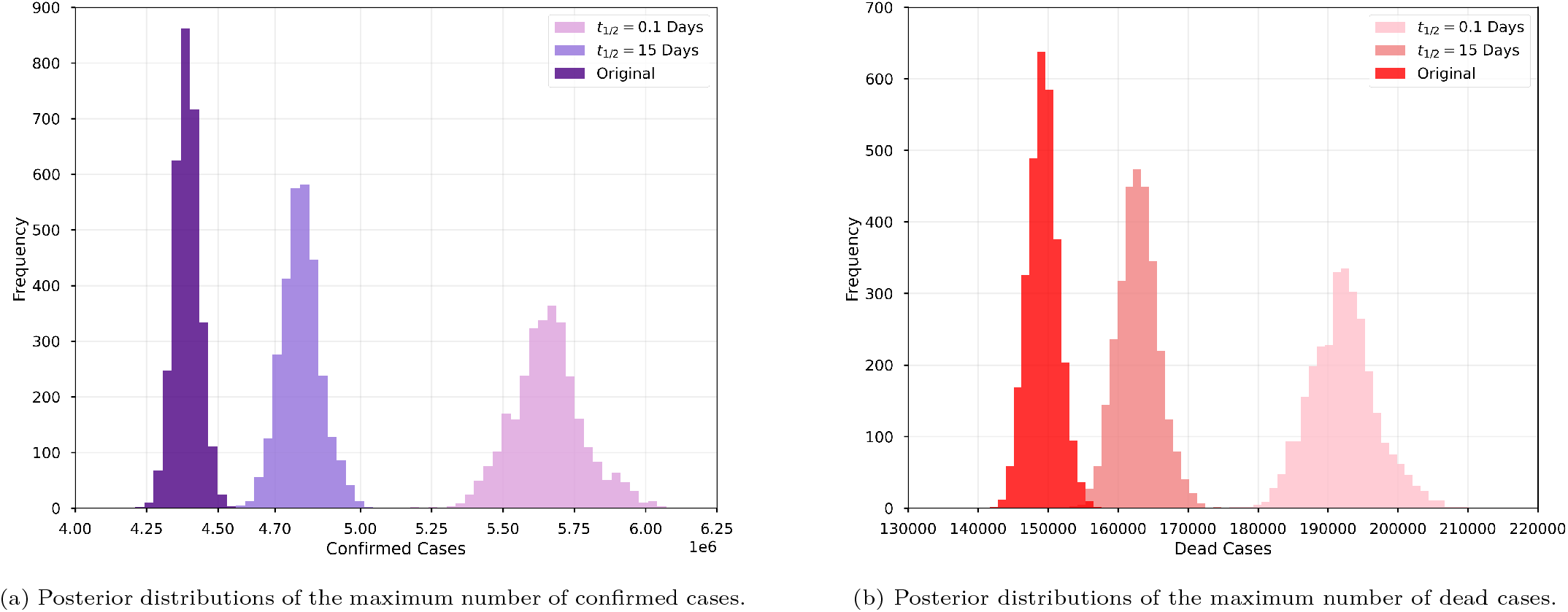
Comparison of the posterior distributions of the maximum numbers of confirmed (a) and dead (b) cases for different social distancing scenarios in BR. The original scenario corresponds to keeping *ω* fixed at its MAP estimate. Analysis is done on the last simulation day (397).

We also remark the importance of the release date of social distancing measures in the following less favorable scenario. Considering that social distancing is released 20 days before the peak of the active cases for BR (*t*_*d*_ = 125 days), disease spreading can yield a critical scenario even when a smooth and gradual release strategy (half-life as *t*_1/2_ = 20 days) is adopted, causing an increase in *C* and *D*, and a drastically longer pandemic period, as shown in Fig. 7. In this case, the peak of active cases would occur on the simulation day 154 (95% CI: 152–158) with around 664.6 thousand (95% CI: 651.5–680.8) active cases, and *D* is expected to be around 270.7 thousand (95% CI: 255.1–288.9) at the end of the simulation, with *C* around 7.976 million (95% CI: 7.542–8.473). Note, however, that the dynamics of *C* and *D* did not reach the stabilization level, reflecting the more critical scenario mentioned earlier.

**Figure 7:**
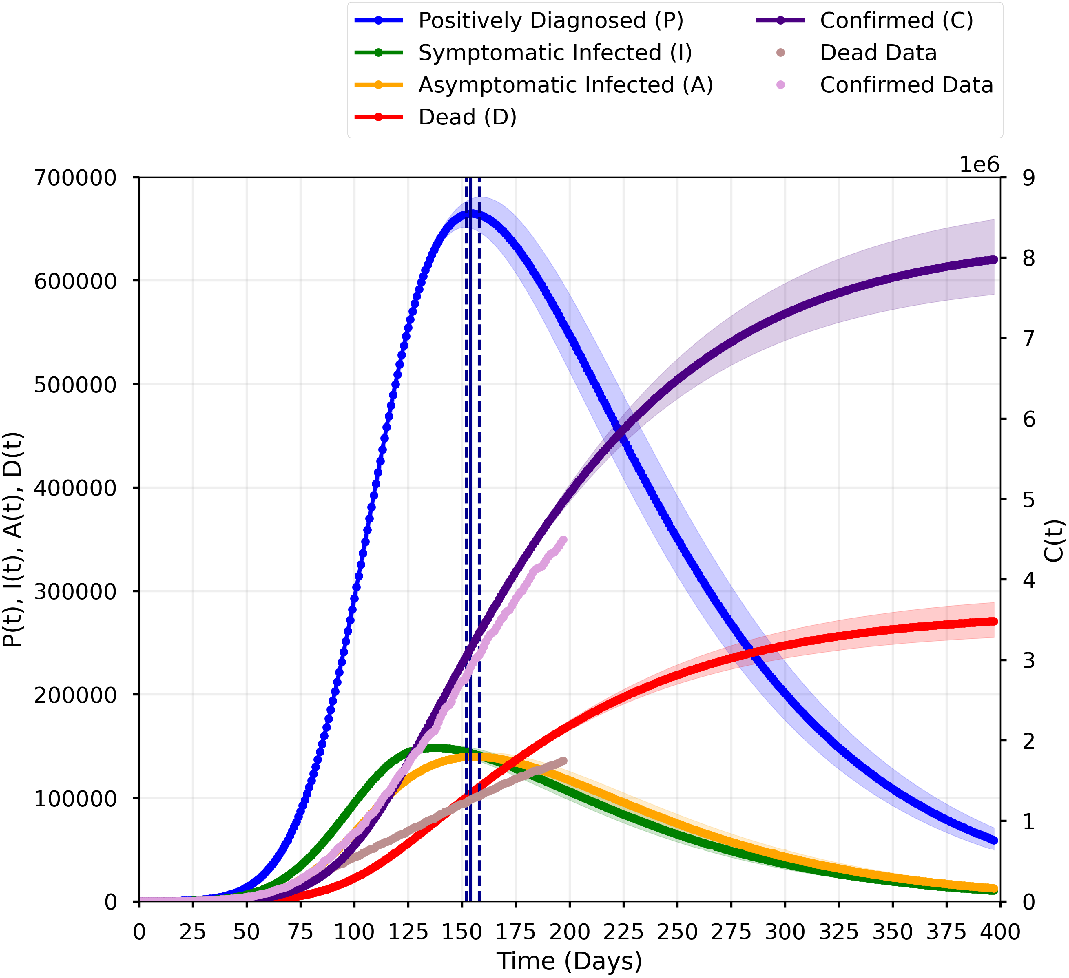
Prediction of *P, I, A, D*, and *C* (right *y*-axis) in BR, considering *t*_1/2_ = 20 days. Social distancing is released 20 days before the peak of active cases (*t*_*d*_ = 125 days).

## 4. Discussion and Conclusions

The present paper contributes to the development of a model framework to investigate the expansion of COVID-19 and the impacts of different measures of social distancing in the presence of uncertainties. We apply the developed approach to model the COVID-19 dynamics in BR and RJ. This high populated Brazilian state was one of the first states to adopt mitigation actions, such as the suspension of classes, cancellation of events, and home isolation [27, 10] (but has not implemented a population quarantine [10]). According to the current data released by the Brazilian Ministry of Health, RJ is one of the most affected states, both in the number of registered cases of COVID-19 and in the number of deaths. For this reason, the present study analyzes the spread in this state, as well as in the country as a whole, in order to assess the particular characteristics of the pandemic at those different spatial scales. Extensive research for the spread of COVID-19 in BR with multiple perspectives has been reported, e.g.,[28, 27, 29, 30, 31, 32]. The present work takes into account factors that are predominant in many underdeveloped countries, such as the current limited testing capacity and the policy to test only severely ill hospitalized individuals. We hope that the present modeling framework brings some insights or guidelines for public health and policy-makers.

Due to data paucity, parameter identifiability is a major difficulty. To overcome this issue, only four model parameters (and two hyperparameters) were calibrated using a Bayesian approach. Other parameters, as well as model initial conditions, were set based on available information on COVID-19 (see the SM for more details). Our simulation forecasts that the peak of active cases occurred on July 28, 2020 (95% CI: 26–29) and July 3, 2020 (95% CI: 2–4) for BR and RJ, respectively. This difference can be explained due to the discrepancy of the spatial scale, and the way the disease has spread along with the different Brazilian locations, considering, for instance, different political measures taken and the social and demographic structure of each Brazilian locality [33]. The social measures implemented in RJ at the beginning of the pandemic seemed to be able to flatten the epidemic curve and postpone the peak of active cases [27, 30]. However, as happened to the whole country [10], these initial social distancing measures were not kept on through the progression of the disease, and that could explain the high number of cases (around 270 thousand confirmed cases and 18 thousand death cases in RJ [34]).

Aiming to evaluate the influence of uncertainties in hard-to-track populations such as undiagnosed infected individuals (*I* and *A*), as well those who carry the disease and are unable to transmit (*E*), we performed a sensitivity analysis to understand which model factors (parameters and initial conditions) play important roles at the various stages of the epidemic for both BR and RJ. The analysis confirms that a proper understanding of how the disease spreads can provide insights and aids to elaborate containment decisions in order to reduce ℛ (*t*). In this sense, considering both BR and RJ, sensitivity analysis suggests that the most influential parameter for a long term perspective is the removal rate parameter (*ω*).

We have shown that the rate at which *S, E, I* and *A* individuals are removed due to social distancing measures significantly affects ℛ(*t*), *C*(*t*) and *D*(*t*). Such average values under the actual social distancing policy are an idealization since actual values are dynamic and spatially heterogeneous, as mentioned in the last paragraph. In order to study the qualitative effects of changes in social distancing policies, we propose to model *ω* as an exponential decay function, which can represent different social distancing relaxation scenarios. When more abrupt social distancing relaxation is implemented after the occurrence of the peak of active cases, it accompanies a longer extension of the duration of the disease, with approximately 29% increase and much higher uncertainty in the projected numbers of *C* and *D* at the end of the simulation. If implemented before the peak, the consequences can be devastating, as indicated by our results. The hypothetical scenario built by considering a slow and gradual release implemented 20 days before the original peak indicates a delay of 9 days in the occurrence of the peak of the active cases, with more than 7.9 million *C* and about 271 thousand *D* accumulated over less than thirteen months of the presence of the disease in BR. Our simulations highlight the importance of relaxing social distancing measures only under a very careful follow-up.

We note that the analysis performed in this paper should be viewed from a qualitative perspective. The conclusions for the considered hypothetical scenarios (with and without social distancing relaxation) are based on model predictions and current employed policies. Model simplifications and the calibration procedure of model parameters can explain potential quantitative discrepancies between our predictions and data. Such simplifications include: (i) homogenization of age, social, and spatial structure, (ii) some model parameters are fixed, with values obtained from the literature, and (iii) model parameters are constant along time (except *ω*, when investigating the impacts of relaxing social distancing measures). Moreover, data have limited information due to sub-notification, since mostly hospitalized cases are tested in BR. These simplifications are inherent of the modeling procedure, and should be viewed as part of the scientific process of the understanding of the natural phenomena.

The present paper can be extended in forthcoming studies, for example, by considering models with spatial heterogeneity, parameter dependence in time and space, and also analyzing data considering sub-notification. Data considering other BR states could also be analyzed employing the same procedure adopted in the present paper. Another possible extension could be related to the determination of limit thresholds in the number of *P* cases under different relaxation social distancing measures.

It is important to highlight the impacts of social distancing relaxation in order to control the COVID-19 pandemic. The adoption of this type of measure directly affects the evolution of ℛ (*t*), as shown in our results. For both BR and RJ, it implies a much slower stagnation or decrease in ℛ (*t*) from the time the measures are implemented. Since ℛ (*t*) is in a controlled situation for both scenarios, i.e. ℛ(*t*) *<* 1, a direct consequence is that the disease would need more time to be eradicated. Our analyses suggest that policies based on short-term social distancing are not enough to control the evolution of the pandemic. If social distancing policy measures are released before the *“optimal time”*, a second peak should be experienced [28]. Some authors argue that longer or even intermittent social distancing will be necessary to avoid recurrent outbreaks. Specifically, [35] examined a range of likely virus transmission scenarios until 2025 and assessed non-pharmaceutical interventions to mitigate the outbreak. They concluded that if the new coronavirus behaves in the same matter to similar viruses we can expect the disease to return in the coming years, depending on the level and duration of immunity, an aspect that remains to be clarified in the future. The discovery of a vaccine, or new treatments, coupled with the testing of the population could alleviate the need for severe social distancing measures to control the disease. Until then, the need to maintain social distancing measures, even if intermittently, must be carefully addressed. Concerning the actual stage of development of vaccines for COVID-19, there are already strategies for the future administration of such vaccines. As an extension of the present paper using our proposed model, one could employ the protocol developed in [36] for the search of the best strategies of vaccine administration. Another potential extension of the present paper is to consider the economic impact of the pandemic in terms of different social distancing strategies adopted by different countries. Our proposed model could be extended in a way to couple the epidemiological and economic aspects, and in this way to be able to have a more complete perspective of the effects of the COVID-19 pandemic [6].

## Data Availability

All related code and data are available at Zenodo.

https://zenodo.org/record/3865730

## Acknowledgments

ACMR, LA, and JVOS would like to thank the support through CNPq projects 132591/2014-6, 301327/2020-3, and 166171/2018-2. This study was financed in part by the Coordenação de Aperfeiçoamento de Pessoal de Nível Superior (CAPES), Finance Code 001.

## Supplementary Material

### SM A. Supplementary Material Information

This Supplementary Material (SM) is organized as follows. Sections SM A.1 and SM A.2 expand the Results section of the main document with additional figures for the BR and RJ scenarios, respectively. The last part of the document shows additional model setting information and the calibration data used in this work.

#### SM A.1. BR: Additional Results

Fig. A.2a shows the prediction of *P, I, A, D*, and *C* in BR. A total of 638.8 thousand (95% CI: 630.6–647.1) active cases are expected on the simulation day 145 (95% CI: 143–146), with *D* and *C* expected to be around 149.3 thousand (95% CI: 145.1–153.6) and 4.392 million (95% CI: 4.306–4.482), respectively. The posterior distribution of the peak position is displayed in Fig. A.2b, with the vertical dashed lines corresponding to those displayed in Fig. A.2a. Fig. A.2c depicts the time evolution of ℛ(*t*) and displays two vertical lines identifying the credible interval (95% CI: 133–136) of the time above which ℛ(*t*) ≤1. The same lines are depicted in Fig. A.2d that shows the uncertainty associated with that time value.

Fig. A.3 provides the model forecasts for the considered hypothetical scenarios of social distancing relaxation. Fig. A.3a shows what to expect in case of a sudden release from social distancing after the simulation day 165 (*t*_1/2_ = 0.1 days as half-life decay). In this case, *C* reaches 5.656 million (95% CI: 5.426–5.930) and *D* 192.2 thousand (95% CI: 183.4–202.4) at the end of the simulation (day 397), which corresponds to an increase of approximately 29% in both values when compared to the original scenario with *ω* fixed at its MAP estimate. For a gradual release after the simulation day 165 (*t*_1/2_ = 15 days as half-life decay), the numbers of *C* and *D* at the end of simulation are 4.790 million (95% CI: 4.660–4.941) and 162.8 thousand (95% CI: 157.3–169.0), respectively, as shown in Fig. A.3b. Figs. A.3c and A.3d compare the variability of *C* and *D* at the end of simulation in the form of box plots for all scenarios considered. Outliers appear as individual points and the samples medians are depicted in red.

#### SM A.2. RJ: Additional Results

Fig. A.5a shows the prediction of *P, I, A, D*, and *C* in RJ. A total of 32.2 thousand (95% CI: 31.7–32.7) active cases are expected on the simulation day 115 (95% CI: 114–116), with *D* and *C* expected to be around 16.6 thousand (95% CI: 16.3–17.0) and 212.5 thousand (95% CI: 207.6–217.7), respectively. The posterior distribution of the peak position is displayed in Fig. A.5b, with the vertical dashed lines corresponding to those displayed in Fig. A.5a. Fig. A.5c depicts the time evolution of ℛ(*t*) and displays two vertical lines identifying the credible interval (95% CI: 103–107) of the time above which ℛ(*t*) ≤1. The same lines are depicted in Fig. A.5d that shows the uncertainty associated with that time value.

Fig. A.6 provides the model forecasts for the considered hypothetical scenarios of social distancing relaxation. Fig. A.6a shows what to expect in case of a sudden release from social distancing after the simulation day 165 (*t*_1/2_ = 0.1 days as half-life decay). In this case, *C* reaches 220.5 thousand (95% CI: 214.4–227.7) and *D* 17.3 thousand (95% CI: 16.9–17.8) at the end of the simulation (day 392), which corresponds to an increase of approximately 4% in both values when compared to the original scenario with *ω* fixed at its MAP estimate. For a gradual release after the simulation day 165 (*t*_1/2_ = 15 days as half-life decay), the numbers of *C* and *D* at the end of simulation are 215.2 thousand (95% CI: 210.0–221.1) and 16.8 thousand (95% CI: 16.5–17.2), respectively, as shown in Fig. A.6b. Figs. A.6c and A.6d compare the variability of *C* and *D* at the end of simulation in the form of box plots for all scenarios considered. Outliers appear as individual points and the samples medians are depicted in red.

---

**Figure A.1:**
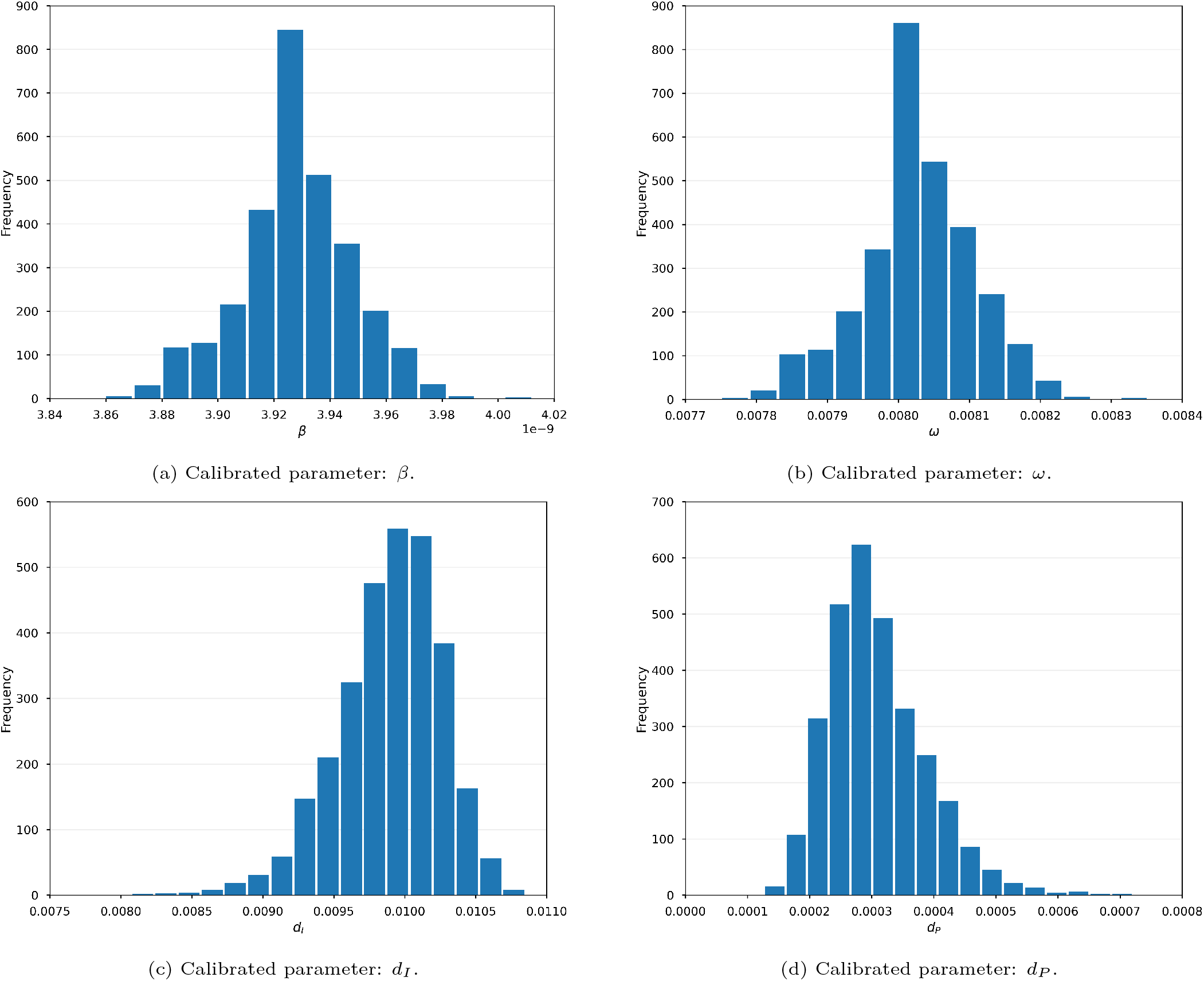
Frequency histograms for the calibrated parameters (BR).

**Figure A.2:**
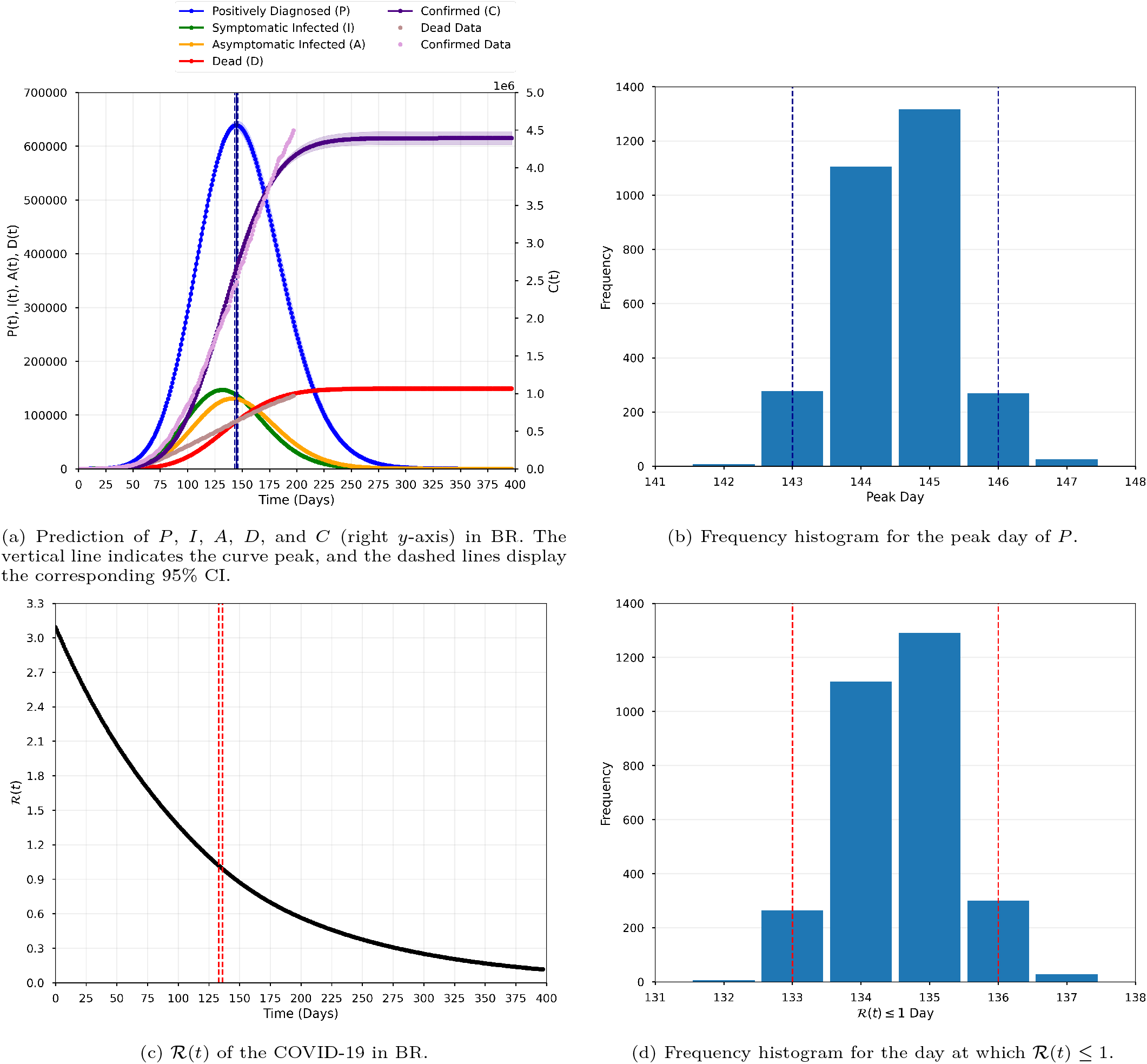
Dynamics of the COVID-19 in BR modeled with the available data (Mathematical Modeling and Methods).

**Figure A.3:**
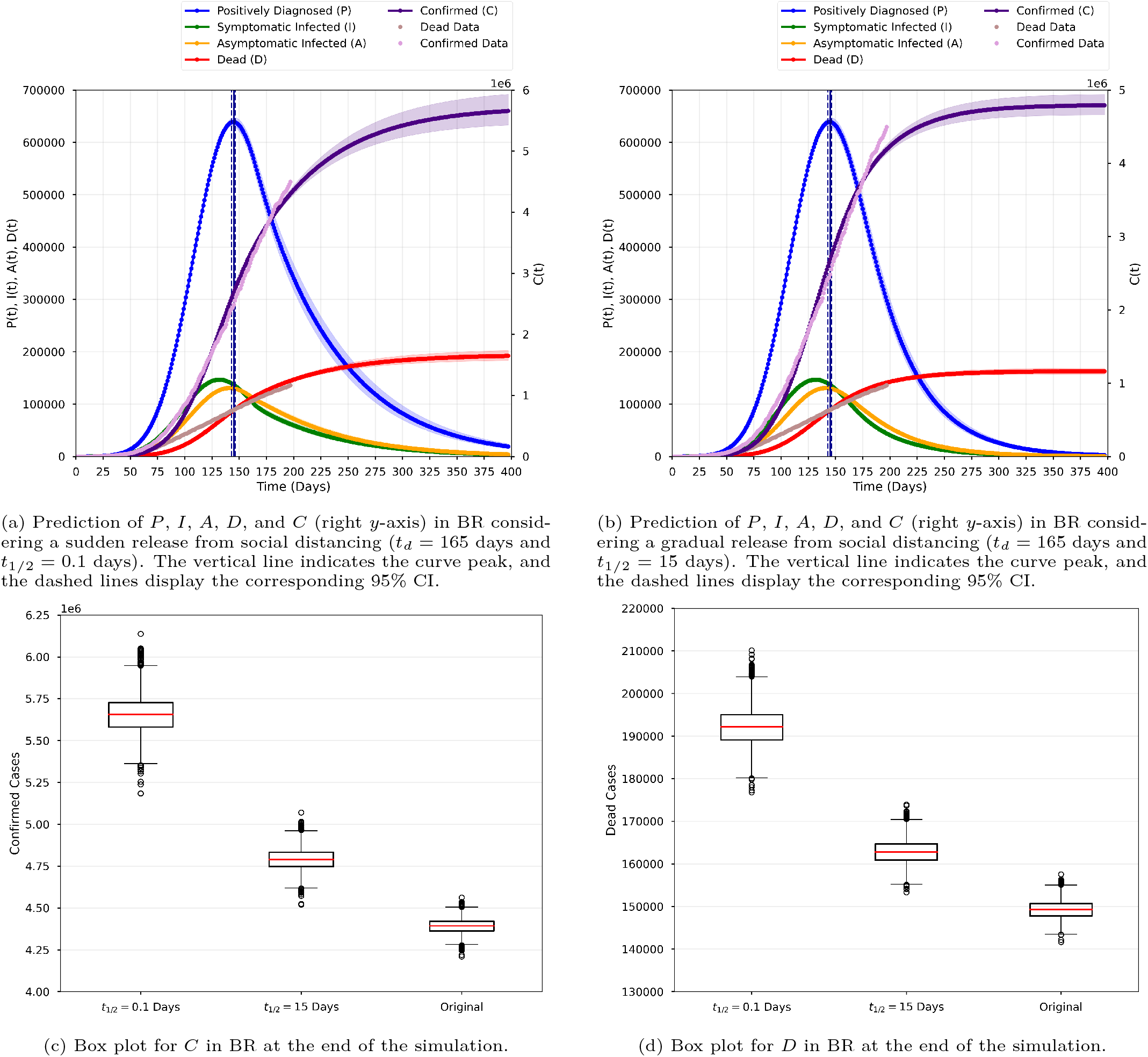
Model forecasts in BR considering sudden and gradual releases from social distancing (*t*_*d*_ = 165 days and *t*_1/2_ = 0.1 and 15 days). The original (baseline) scenario of the main text is also presented to ease comparison.

**Figure A.4:**
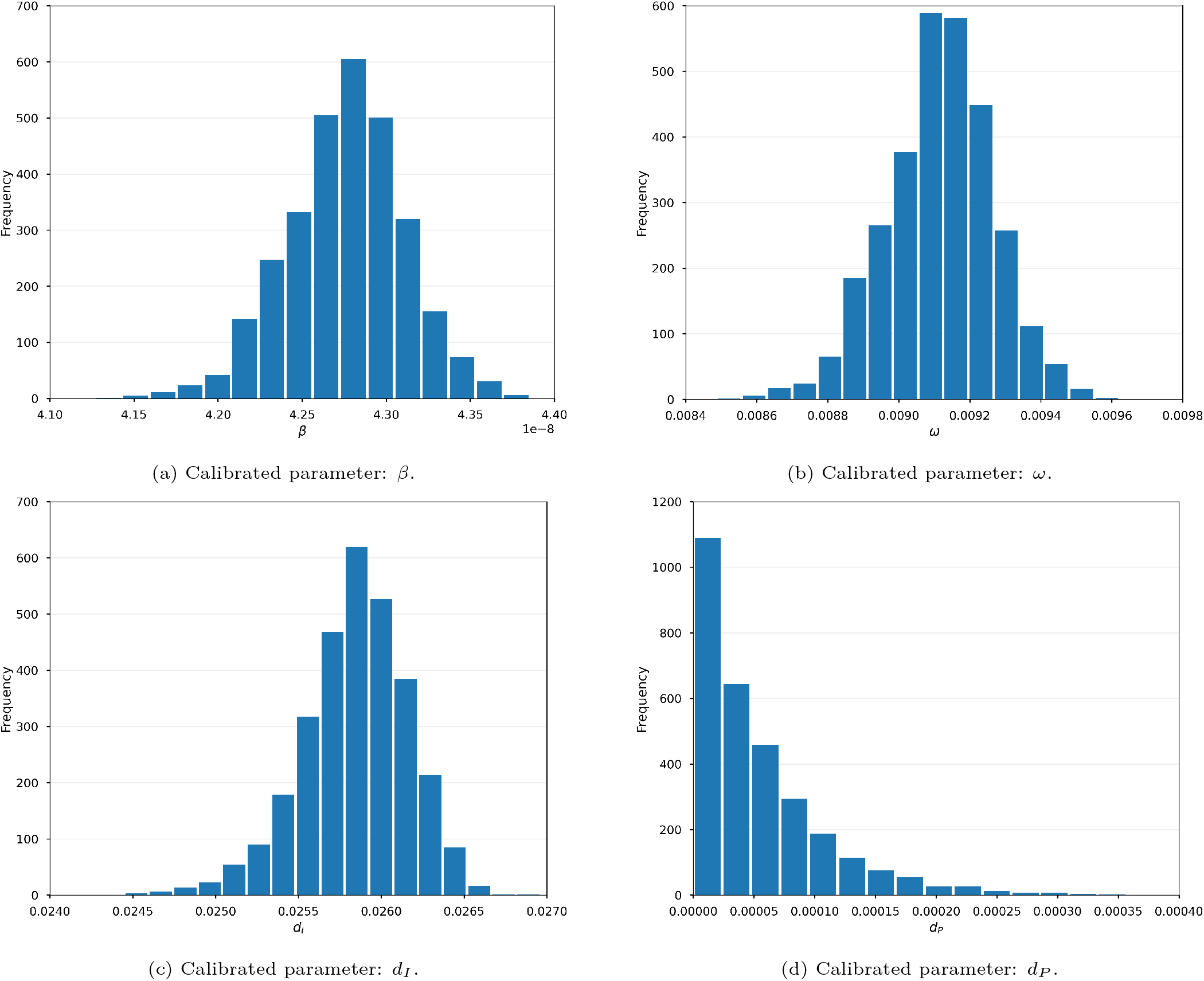
Frequency histograms for the calibrated parameters (RJ).

**Figure A.5:**
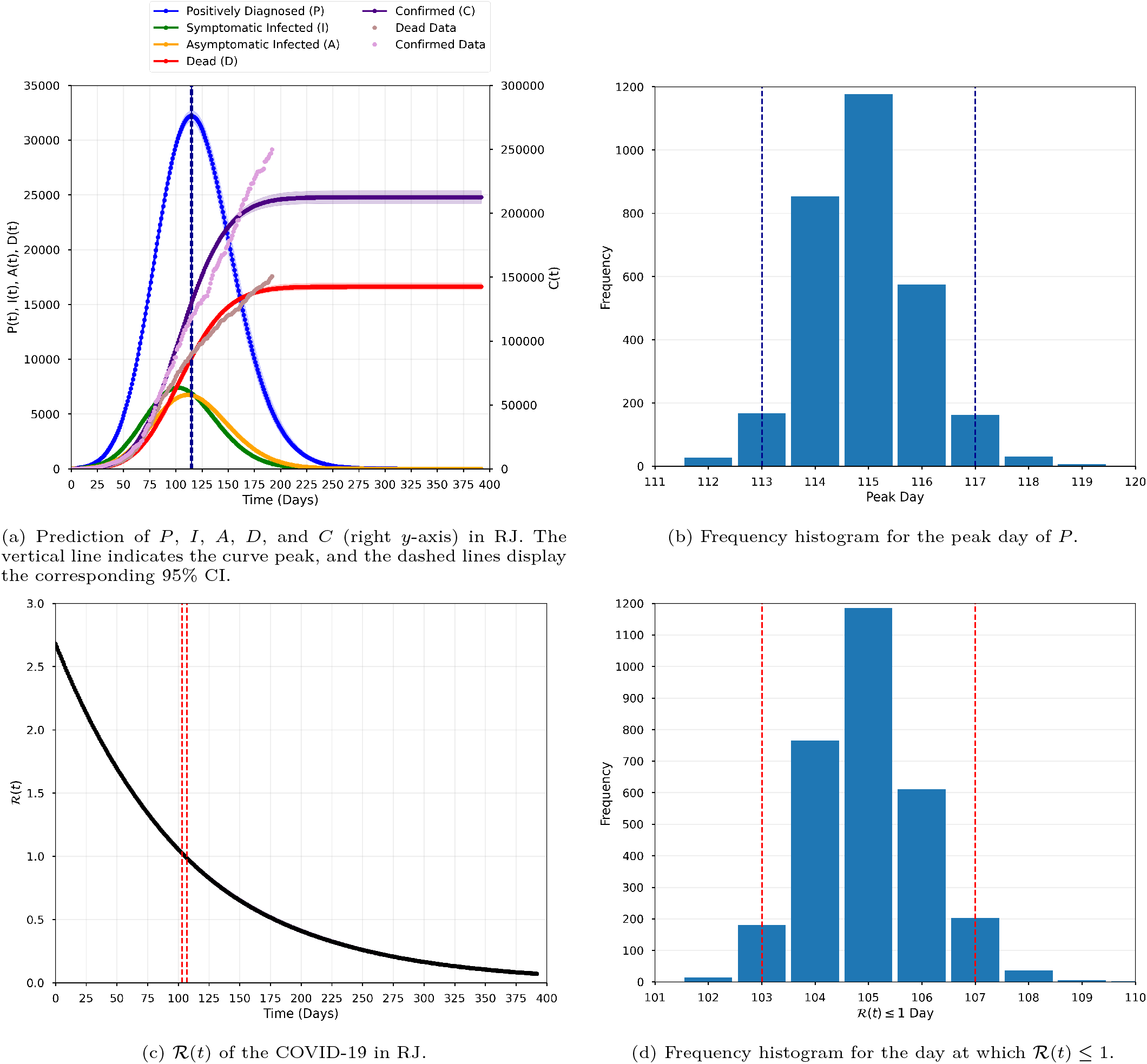
Dynamics of the COVID-19 in RJ modeled with the available data (Mathematical Modeling and Methods).

**Figure A.6:**
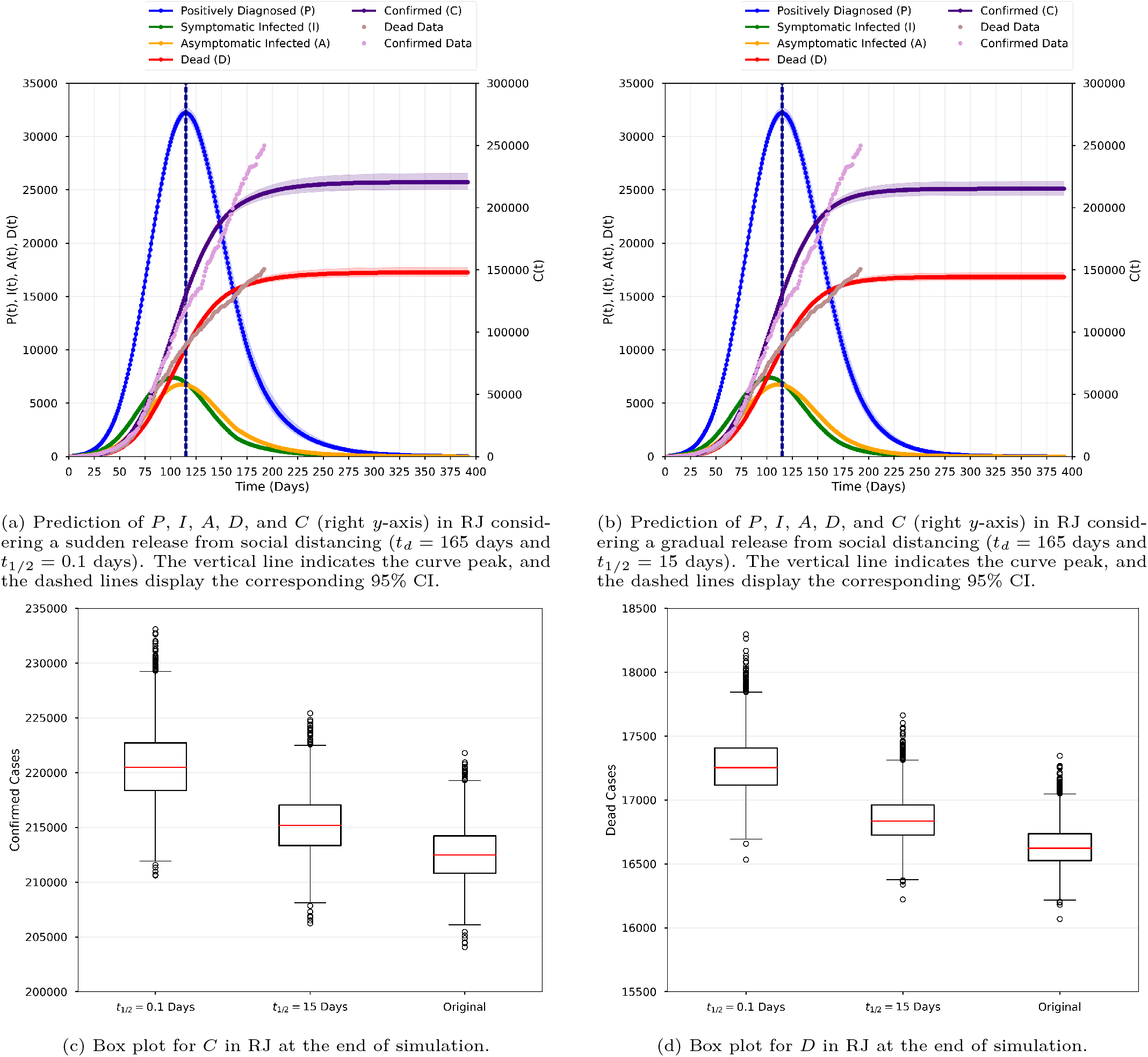
Model forecasts in RJ considering sudden and gradual releases from social distancing (*t*_*d*_ = 165 days and *t*_1/2_ = 0.1 and 15 days). The original (baseline) scenario of the main text is also presented to ease comparison.

**Table A.1:** Model parameters.

| Parameter | Meaning |
| --- | --- |
| $\beta$ | Rate of transmission per contact with symptomatic infected individual |
| $\mu$ | Rate of transmission per contact with asymptomatic infected individual |
| $\sigma$ | Transition rate from exposed to infected individuals |
| $\rho$ | Proportion of infected individuals who have symptoms |
| $\varepsilon_I$ | Diagnosis rate of hospitalized individuals |
| $\gamma_A$ | Recovery rate of asymptomatic infected individuals |
| $\gamma_I$ | Recovery rate of symptomatic infected individuals |
| $\gamma_P$ | Recovery rate of hospitalized individuals |
| $d_I$ | Death rate of hospitalized individuals |
| $d_P$ | Death rate of symptomatic infected individuals |
| $\omega$ | The rate at which susceptible, exposed, and infected individuals are removed due to social distancing measures |

**Table A.2:**
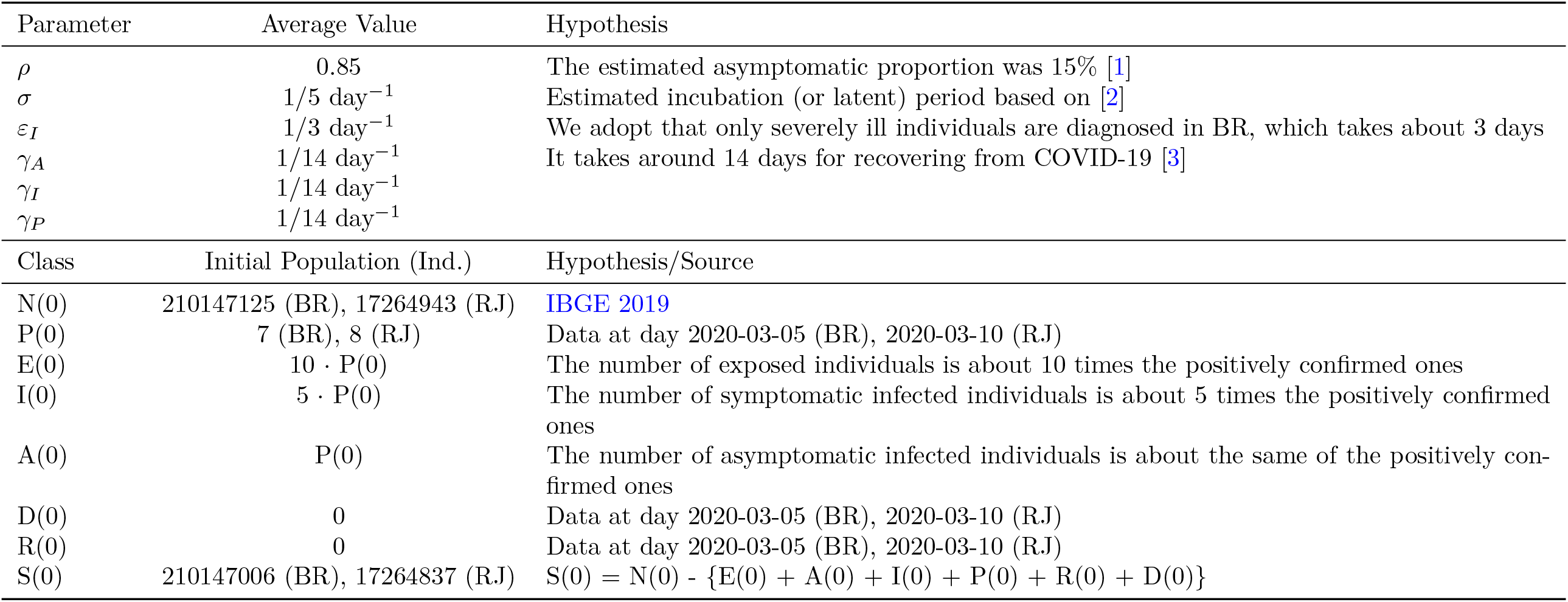
Fixed values for estimated parameters and ICs.

## References

[1] World Health Organization, Coronavirus disease 2019 (COVID-19): Weekly epidemiological update, www.who.int/emergencies/diseases/novel-coronavirus-2019 (Accessed September 30, 2020).

[2] M. Mesel-Lemoine, J. Millet, P. O. Vidalain, H. Law, A. Vabret, V. Lorin, N. Escriou, M. L. Albert, B. Nal, F. Tangy, A human coronavirus responsible for the common cold massively kills dendritic cells but not monocytes, Journal of Virology 86 (14) (2012) 7577–7587. doi:10.1128/JVI.00269-12.

[3] Worldometer, Coronavirus, https://www.worldometers.info/coronavirus/ (Accessed April 20, 2020).

[4] J. R. Koo, A. R. Cook, M. Park, Y. Sun, H. Sun, J. T. Lim, C. Tam, B. L. Dickens, Interventions to mitigate early spread of SARS-CoV-2 in Singapore: a modelling study, The Lancet Infectious Diseases.doi:10.1016/S1473-3099(20)30162-6.

[5] C. T. Bauch, J. O. Lloyd-Smith, M. P. Coffee, A. P. Galvani, Dynamically modeling SARS and other newly emerging respiratory illnesses: past, present, and future, Epidemiology 16 (6) (2005) 791–801. doi:10.1097/01.ede.0000181633.80269.4c.

[6] J. Wang, Mathematical models for covid-19: Applications, limitations, and potentials, Journal of Public Health and Emergency 4. doi:10.21037/jphe-2020-05.

[7] F. A. L. Marson, COVID-19–6 million cases worldwide and an overview of the diagnosis in Brazil: A tragedy to be announced, Diagnostic Microbiology and Infectious Disease 98 (2) (2020) 115113. doi:10.1016/j.diagmicrobio.2020.115113.

[8] B. F. Maier, D. Brockmann, Effective containment explains subexponential growth in recent confirmed COVID-19 cases in China, Science 368 (6492) (2020) 742–746. doi:10.1126/science.abb4557.

[9] Decreto 46980 de 19/03/2020, Medidas de enfrentamento, Governo RJ (in Portuguese) (Accessed April 20, 2020).

[10] L. L. S. Silva, A. F. R. Lima, D. A. Polli, P. F. S. Razia, L. F. A. Pavão, M. A. F. H. Cavalcanti, C. M. Toscano, Social distancing measures in the fight against COVID-19 in Brazil: description and epidemiological analysis by state, Cadernos de Saúde Pública 36 (2020) e00185020. doi:10.1590/0102-311X00185020.

[11] O. Diekmann, J. A. P. Heesterbeek, M. G. Roberts, The construction of next-generation matrices for compartmental epidemic models, Journal of The Royal Society Interface 7 (2010) 873–885. doi:10.1098/rsif.2009.0386.

[12] A. Tarantola, Inverse problem theory and methods for model parameter estimation, SIAM, Philadelphia, 2005. doi:10.1137/1.9780898717921.

[13] J. Salvatier, T. V. Wiecki, C. Fonnesbeck, Probabilistic programming in Python using PyMC3, PeerJ Computer Science 2 (2016) e55. doi:10.7717/peerj-cs.55.

[14] S. E. Minson, M. Simons, J. L. Beck, Bayesian inversion for finite fault earthquake source models I – theory and algorithm, Geophysical Journal International 194 (3) (2013) 1701–1726. doi:10.1093/gji/ggt180.

[15] D. Volpatto, A. C. M. Resende, L. Anjos, J. V. O. Silva, C. M. Dias, R. C. Almeida, S. M. C. Malta, pydemic: Scripts for BR/RJ social distancing study (May 2020). doi:10.5281/zenodo.3865730.

[16] L. Petzold, Automatic selection of methods for solving stiff and nonstiff systems of ordinary differential equations, SIAM Journal on Scientific and Statistical Computing 4 (1) (1983) 136–148. doi:10.1137/0904010.

[17] P. Virtanen, R. Gommers, T. E. Oliphant, M. Haberland, T. Reddy, D. Cournapeau, E. Burovski, P. Peterson, W. Weckesser, et al., SciPy 1.0: Fundamental Algorithms for Scientific Computing in Python, Nature Methods 17 (2020) 261–272. doi:10.1038/s41592-019-0686-2.

[18] A. C. Hindmarsh, ODEPACK, a systematized collection of ODE solvers, Scientific Computing 1 (1983) 55–64.

[19] Brazilian Health Ministry, Coronavirus panel (in Portuguese), https://covid.saude.gov.br/ (Accessed Sep 18, 2020).

[20] F. Campolongo, J. Cariboni, A. Saltelli, An effective screening design for sensitivity analysis of large models, Environmental Modelling & Software 22 (10) (2007) 1509–1518. doi:10.1016/j.envsoft.2006.10.004.

[21] J. Herman, W. Usher, SALib: an open-source Python library for sensitivity analysis, Journal of Open Source Software 2 (9) (2017) 97. doi:10.21105/joss.00097.

[22] G. A. K. Van Voorn, B. W. Kooi, Combining bifurcation and sensitivity analysis for ecological models, The European Physical Journal Special Topics 226 (9) (2017) 2101–2118. doi:10.1140/epjst/e2017-70030-2.

[23] S. A. Lauer, K. H. Grantz, Q. Bi, F. K. Jones, Q. Zheng, H. R. Meredith, A. S. Azman, N. G. Reich, J. Lessler, The incubation period of coronavirus disease 2019 (COVID-19) from publicly reported confirmed cases: estimation and application, Annals of Internal Medicine 172 (9) (2020) 577–582. doi:10.7326/M20-0504.

[24] K. Mizumoto, K. Kagaya, A. Zarebski, G. Chowell, Estimating the asymptomatic proportion of coronavirus disease 2019 (COVID-19) cases on board the Diamond Princess cruise ship, Yokohama, Japan, 2020, Eurosurveillance 25 (10) (2020) 2000180. doi:10.2807/1560-7917.

[25] F. Pan, T. Ye, P. Sun, S. Gui, B. Liang, L. Li, D. Zheng, J. Wang, R. L. Hesketh, L. Yang, et al., Time course of lung changes on chest ct during recovery from 2019 novel coronavirus (COVID-19) pneumonia, Radiology (2020) 200370 doi:10.1148/radiol.2020200370.

[26] P. Van den Driessche, J. Watmough, Reproduction numbers and sub-threshold endemic equilibria for compartmental models of disease transmission, Mathematical Biosciences 180 (1-2) (2002) 29–48. doi:10.1016/S0025-5564(02)00108-6.

[27] N. Crokidakis, COVID-19 spreading in Rio de Janeiro, Brazil: do the policies of social isolation really work?, medRxiv (2020) medRxiv:2020.04.27.20081737 doi:10.1101/2020.04.27.20081737.

[28] S. B. Bastos, D. O. Cajueiro, Modeling and forecasting the early evolution of the COVID-19 pandemic in Brazil, arXiv e-prints (2020) 2003.14288.

[29] N. Crokidakis, Modeling the early evolution of the COVID-19 in Brazil: results from a Susceptible-Infectious-Quarantined-Recovered (SIQR) model, International Journal of Modern Physics Cdoi:10.1142/S0129183120501351.

[30] N. Crokidakis, COVID-19 spreading in Rio de Janeiro, Brazil: do the policies of social isolation really work?, Chaos, Solitons & Fractals (2020) 109930 doi:10.1016/j.chaos.2020.109930.

[31] R. A. Schulz, C. H. Coimbra-Araújo, S. W. S. Costiche, COVID-19: A model for studying the evolution of contamination in Brazil, arXiv e-prints (2020) 2003.13932.

[32] R. F. Reis, B. M. Quintela, J. O. Campos, J. M. Gomes, B. M. Rocha, M. Lobosco, R. W. dos Santos, Characterization of the COVID-19 pandemic and the impact of uncertainties, mitigation strategies, and underreporting of cases in South Korea, Italy, and Brazil, Chaos, Solitons & Fractals (2020) 109888 doi:10.1016/j.chaos.2020.109888.

[33] F. A. L. Marson, M. M. Ortega, COVID-19 in Brazil, Pulmonology 26 (4) (2020) 241–244. doi:10.1016/j.pulmoe.2020.04.008.

[34] Governo do Rio de Janeiro, Boletim epidemiológico diário estado do Rio de Janeiro (in Portuguese), https://coronavirus.rj.gov.br/boletins/ (Accessed October 2, 2020).

[35] S. M. Kissler, C. Tedijanto, E. Goldstein, Y. H. Grad, M. Lipsitch, Projecting the transmission dynamics of SARS-CoV-2 through the postpandemic period, Science 368 (6493) (2020) 860–868. doi:10.1126/science.abb5793.

[36] G. B. Libotte, F. S. Lobato, A. J. S. Neto, G. M. Platt, Determination of an optimal control strategy for vaccine administration in COVID-19 pandemic treatment, Computer Methods and Programs in Biomedicine 196 (2020) 105664. doi:10.1016/j.cmpb.2020.105664.

## References

[1] K. Mizumoto, K. Kagaya, A. Zarebski, G. Chowell, Estimating the asymptomatic proportion of coronavirus disease 2019 (COVID-19) cases on board the Diamond Princess cruise ship, Yokohama, Japan, 2020, Eurosurveillance 25 (10) (2020) 2000180. doi:10.2807/1560-7917.

[2] S. A. Lauer, K. H. Grantz, Q. Bi, F. K. Jones, Q. Zheng, H. R. Meredith, A. S. Azman, N. G. Reich, J. Lessler, The incubation period of coronavirus disease 2019 (COVID-19) from publicly reported confirmed cases: estimation and application, Annals of Internal Medicine 172 (9) (2020) 577–582. doi:10.7326/M20-0504.

[3] F. Pan, T. Ye, P. Sun, S. Gui, B. Liang, L. Li, D. Zheng, J. Wang, R. L. Hesketh, L. Yang, et al., Time course of lung changes on chest ct during recovery from 2019 novel coronavirus (COVID-19) pneumonia, Radiology (2020) 200370 doi: 10.1148/radiol.2020200370.

